# A meta-analysis on the role of children in SARS-CoV-2 in household transmission clusters

**DOI:** 10.1101/2020.03.26.20044826

**Authors:** Yanshan Zhu, Conor J. Bloxham, Katina D. Hulme, Jane E. Sinclair, Zhen Wei Marcus Tong, Lauren E. Steele, Ellesandra C. Noye, Jiahai Lu, Yao Xia, Keng Yih Chew, Janessa Pickering, Charles Gilks, Asha C. Bowen, Kirsty R Short

**Affiliations:** School of Chemistry and Molecular Biosciences, The University of Queensland, Brisbane Australia; School of Biomedical Science, The University of Queensland, Brisbane, Australia; School of Public Health, Sun Yat-sen University, Zhongshan 2nd Road, Guangzhou 510080, Guangdong, China; School of Science, Edith Cowan University, Australia; School of Biomedical Science, The University of Western Australia, Perth, Australia; Wesfarmer’s Centre for Vaccines and Infectious diseases, Telethon Kids Institute, University of Western Australia, Nedlands, Perth, Western Australia; Department of Infectious Diseases, Perth Children’s Hospital, Nedlands, Perth, Western Australia; School of Public Health, The University of Queensland, Brisbane Australia; Australian Infectious Diseases Research Centre, The University of Queensland, Brisbane, Australia

## Abstract

The role of children in the spread of SARS-CoV-2 remains highly controversial. To address this issue, we performed a meta-analysis of the published literature on household SARS-CoV-2 transmission clusters (n=213 from 12 countries). Only 8 (3.8%) transmission clusters were identified as having a paediatric index case. Asymptomatic index cases were associated with a lower secondary attack in contacts than symptomatic index cases (estimate risk ratio [RR], 0.17; 95% confidence interval [CI], 0.09-0.29). To determine the susceptibility of children to household infections the secondary attack rate (SAR) in paediatric household contacts was assessed. The secondary attack rate in paediatric household contacts was lower than in adult household contacts (RR, 0.62; 95% CI, 0.42-0.91). These data have important implications for the ongoing management of the COVID-19 pandemic, including potential vaccine prioritization strategies.

**40-word summary:** In household transmission clusters of SARS-CoV-2 children are unlikely to be the index case. Children are also less likely than adults to be infected with SARS-CoV-2 from a family member.

## INTRODUCTION

At the time of writing severe acute respiratory syndrome coronavirus 2 (SARS-CoV-2) has infected >50 million people resulting in >1 million deaths [1].Large data analyses have shown that the elderly are particularly susceptible to severe forms of coronavirus disease 2019 (COVID-19) [2]. However, the role of children in the transmission of SARS-CoV-2 remains controversial [3-9]. During a typical influenza virus season, children have been identified as having the highest infection rate of any age group (up to 43%). Accordingly, children may play a major role in the spread of influenza virus and are a key target population for influenza vaccination to prevent infection and reduce transmission[10]. In the context of coronaviruses, paediatric infections with SARS-CoV-1, SARS-CoV-2 and MERS are typically mild [9, 10]. Nevertheless, a lower incidence of clinical symptoms raises concerns that children could be an important, undetected source of SARS-CoV-2 in transmission in the community [8, 11]. Answering this question is of key importance to public health as it will help identify priority groups for vaccination. However, findings remain controversial, with some studies suggesting that children may play a key role in disease transmission and shed virus at equivalent titres to adults [12-17]. In contrast, others find little evidence of paediatric infections or spread [7, 8, 18-21]. Moreover, it is unclear if SARS-CoV-2 transmission differs amongst children of differing age groups.

Studying the source and route of viral transmission from children in the community is fraught with difficulties, due to the multiple different potential sources of infection. Furthermore, it is thought that households are one of the most common settings in SARS-CoV-2 transmission [22]. Household transmission clusters therefore offer the unique opportunity to study viral transmission and susceptibility to infection in a more defined setting.

To address the role of children in the transmission of SARS-CoV-2, we performed a meta-analysis on household transmission clusters. We investigated prevalence of paediatric index cases in household transmission clusters of SARS-CoV-2 as well as the secondary attack rate (SAR) of different age groups.

## METHODS

### Definitions

A household transmission cluster was defined as a group of ≥ 2 confirmed cases of SARS-CoV-2 infections in co-habiting individuals in whom the diagnosis of cases occurred within 2 weeks of each other. The index case was defined as the individual in the household cluster who first developed symptoms. Household contacts were defined as co-habiting individuals, typically family members, close relatives, housemates or house helpers. An individual with laboratory confirmation of SARS-CoV-2 was considered to be infected. Household secondary attack rates were defined as the proportion of confirmed infections among all household contacts. Unless otherwise stated, adults were defined as individuals ≥ 18 years whilst children were defined as individuals <18 years of age.

### Data collection

Following the PRISMA statement for the reporting of meta-analysis [23], we searched published, de-identified, data made available between Dec 1, 2019, and August 24, 2020. Information was accessed from the World Health Organization news^11^, Google Scholar, PubMed, the Lancet COVID-19 resource centre^12^, Clinical Infectious Disease Journal and New England Journal of Medicine. We searched for databases using the search terms (“COVID-19” OR “SARS-CoV-2” OR) AND (“household transmission” OR “family cluster” OR “household contact”) AND (“transmissibility” OR “attack rate”). To identify missing studies, we checked the reference list for each selected paper. Studies that were duplicate publications, pre-prints and/or reviews were excluded (Figure 1).

**Figure 1.**
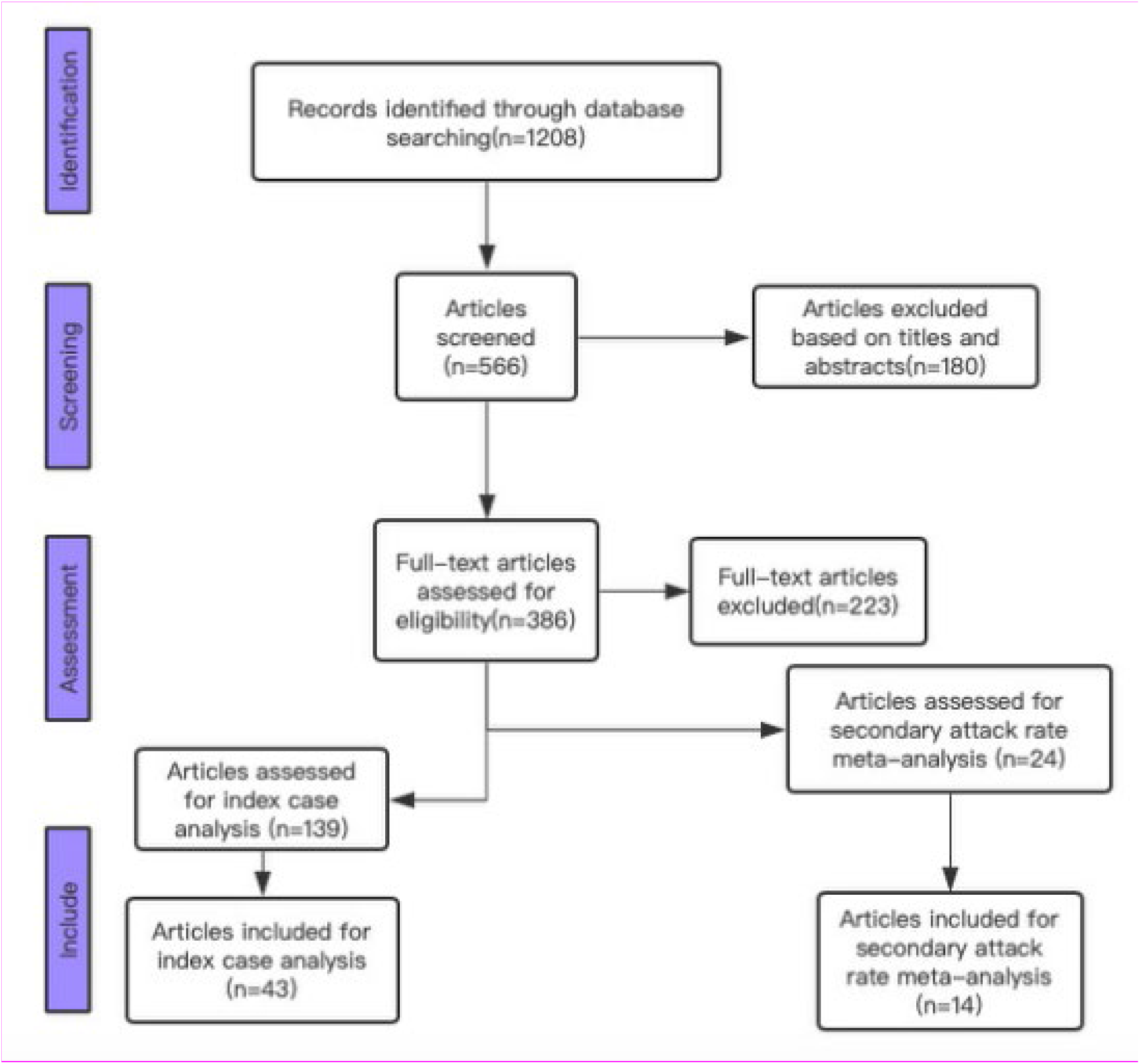
Preferred Reporting Items for Systematic Reviews and Meta-analysis (PRISMA) flow diagram.

Our search strategy aimed to identify all articles that assessed the prevalence of children as index cases in a family SARS-CoV-2 cluster and/or the secondary attack rate of children and adults in household transmission clusters. Depending on the level of information available, studies were included in the index case analysis or the secondary attack rate meta-analysis.

All studies included in the index case analysis were household SARS-CoV-2 transmission clusters that i) identified the index case of the cluster ii) defined the number of infected contacts in the household and iii) recorded the initial disease onset date of all cases in the cluster.

All studies included in the secondary attack rate meta-analysis were household SARS-CoV-2 transmission clusters that i) defined the secondary attack rate within the cluster and ii) defined the age of contact cases in the cluster. Studies which did not meet bare minimum data required for the index case analysis nor the secondary attack rate meta-analysis were excluded (Figure 1). Where the same family cluster was included in more than published report, data was only extracted from one study. Collected data was verified by a second researcher.

### Statistical analysis

Susceptibility to infection was estimated by calculating the secondary attack rate for household close contacts associated with the index case in each transmission cluster. We estimated the Relative Risk (RRs) for SARS CoV-2 infection stratified by the age of household contacts for each study. We then pooled these RRs using a random effects model with DerSimonian and Laird weights[24]. We used a random effects model, equalizing the weight of the studies to the pooled estimate. Where relevant, we stratified the analysis by pre-specified characteristics. 95% confidence intervals (CI) were used to assess statistical significance in all models. The I^2^ statistic was used to evaluate heterogeneity between studies. A threshold of I^2^>50% was used as indicating statistically significant heterogeneity. All summary analyses and meta-analysis were performed using R statistical software (version 3.6.1).

## RESULTS

We identified 1208 articles that described SARS-CoV-2 household transmission clusters, rejected 1151 articles due to a lack of sufficient and or appropriate data and derived a total of 57 articles. Household transmission clusters were drawn from cases in 12 countries: China, Japan, France, Germany, Italy, USA, Vietnam, Malaysia, Singapore, Morocco, Greece and South Korea. 43 articles were included in the index case analysis[5, 25-66] whilst 14 articles were used in the secondary attack rate meta-analysis[66-79]. The full detail of all family clusters and characteristics of studies included in meta-analysis are shown in Supplementary Tables 1 and 2.

### Children are infrequently identified as the index case of household SARS-CoV-2 clusters

In analysis of the cluster index cases, we included 43 articles, in which there were 213 SARS-CoV-2 transmission clusters and only 3.8% (8/213) were identified as having a paediatric index case (Table 1 & Supplementary Tables 1 & 3). Of 611 individuals in the 213 clusters there were 102 children. These paediatric cases only caused 4.0% (16/398) of all secondary cases, compared to the 97.8% of secondary cases that occurred when an adult was identified as the index case in the cluster (Table 1).

**Table 1.** Household transmission clusters of SARS-CoV-2 stratified by the age of the index case.

| Characteristics | SARS-CoV-2 household transmission cases |  |
| --- | --- | --- |
|  | Cluster (n=213),<br>No. (%) | Secondary cases (n=398), No. (%) |
| Child as the index case | 8 (3.8) | 16 (4.0) |
| Adult as the index case | 205 (96.2) | 382 (96.0) |

The limited number of defined SARS-CoV-2 household clusters with children as the index case could have been influenced by the fact that COVID-19 in children is frequently asymptomatic [11]. Accordingly, it is possible that within a household cluster, children were not correctly identified as the index case of the infection (i.e. the first to develop symptoms) and were instead mistakenly identified as a contact case. To exclude this possibility, we reanalysed the data looking at household clusters where a paediatric contact case was SARS-CoV-2 positive but asymptomatic. In such a scenario we assumed the child to be the ‘true’ index case of the cluster. Clusters where the asymptomatic/symptomatic status of the contact cases was not described were excluded from the analysis. Even with this broader definition, only 39 (18.5%) children were identified as the index case in the household clusters (Table 2).

**Table 2.** Household transmission clusters of SARS-CoV-2 where any asymptomatic, SARS-CoV-2 positive children are assumed to be the index case of the cluster.

| Characteristics | SARS-CoV-2 household transmission cases |  |
| --- | --- | --- |
|  | Cluster (n=211), No. (%) | Secondary cases (n=395), No. (%) |
| Child as the index case<br>(including asymptomatic children) | 39 (18.5) | 80 (21.7) |
| Adult as the index case | 172 (81.5) | 315 (79.7) |

It is also possible that these data were influenced by the fact that early in the SARS-CoV-2 outbreak, infections were associated with travel to outbreak areas (i.e. initially to Wuhan itself and later to the entirety of Hubei). Travel is much more likely to be undertaken by an adult in the family, potentially cofounding the results. To control for this factor, we reanalysed the data only including household transmission clusters where the index case had no history of travel or the whole family was associated with an outbreak area. Clusters where this information was not available were excluded from the analysis. This resulted in a total 152 index cases, 32 of which (21.1%) were identified as having a child as the index case in the cluster (Table 3).

**Table 3.** Household transmission clusters of SARS-CoV-2 accounting for the travel of adults to outbreak areas.

| Characteristics | SARS-CoV-2 household transmission cases |  |
| --- | --- | --- |
|  | Cluster (n=152),<br>No. (%) | Secondary cases (n=264),<br>No. (%) |
| Child as the index case | 32 (21.1) | 68 (25.8) |
| Adult as the index case | 120 (78.9) | 196 (74.2) |

A final factor which may have confounded this analysis is the fact that in some countries a strict lockdown was imposed during the period of data collection. This would have limited the activity of children outside of the house and may therefore have artificially reduced the number of children identified as an index case. To control for this factor, a sub-analysis was performed only using data collected when the regional area or country was not in a period of ‘lockdown’. In this sub-analysis only 3.0% of clusters were associated with a paediatric index case (Table 4, Supplementary Table 1).

**Table 4.** Household transmission clusters of SARS-CoV-2 in the absence of regional or national ‘lockdown’.

| Characteristics | SARS-CoV-2 household transmission cases |  |
| --- | --- | --- |
|  | Cluster (n=199),<br>No. (%) | Secondary cases (n=366),<br>No. (%) |
| Child as the index case | 6 (3.0) | 12 (3.3) |
| Adult as the index case | 193 (97.0) | 354 (96.7) |

### Asymptomatic index cases are associated with a lower secondary attack rate

We then further examined the household clusters identified in Table 1 where a child was identified as the index case in order to define the secondary attack rate of cohabiting family members (Supplementary Table 4). Clusters where the total number of infected and uninfected family members was not recorded was excluded from the analysis. Accordingly, sufficient information was only available to calculate the secondary attack rate of five clusters with a paediatric index case (mean = 46.7%; SD = 28.2%). Three of these index cases were < 10 years and 3 index cases were aged 10 – 19 years. Only 22 clusters with an adult index case were eligible for this analysis (mean = 65.8%; SD = 23.3%) (Supplementary Table 5). Therefore, there was insufficient case numbers to determine whether children are more or less able to transmit SARS-CoV-2 in a household setting compared to adult index cases.

It has previously been suggested that asymptomatic individuals may be less infectious than those who develop symptoms [7, 80, 81]. The prevalence of mild/asymptomatic infections in children may therefore affect the secondary attack rate in SARS-CoV-2 household clusters where children were identified as the index case. To assess this possibility, we examined the secondary attack rate in household clusters where the index case was symptomatic vs. the secondary attack rate in household clusters where the index case was asymptomatic, (but known to be SARS-CoV-2 positive Figure 2). Asymptomatic index cases were associated with a significantly lower secondary attack in contacts than symptomatic index cases RR = 0.17 (95% CI, 0.09-0.29), although a significant overall effect was observed with heterogeneity (I^2^=87%, P<0.01).

**Figure 2.**
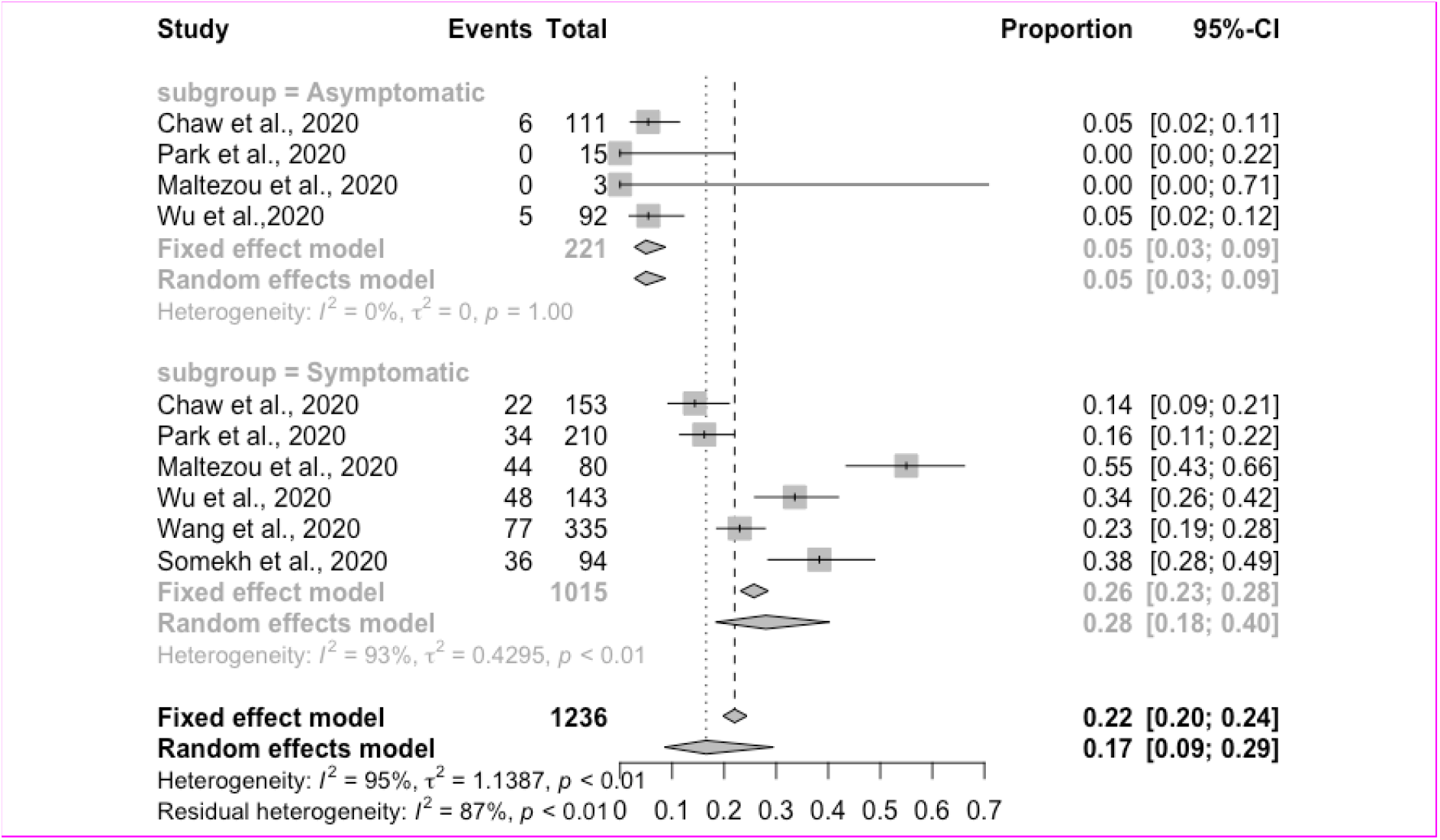
The secondary attack rate of household transmission stratified by SARS-CoV-2 positive symptomatic and asymptomatic index cases.

### Children have a lower secondary attack rate than adults in household SARS-CoV-2 transmission clusters

Several studies have suggested that children are less likely than adults to be infected with SARS-CoV-2 [20, 82]. However, small scale studies can be biased by the fact that community testing is often only performed on symptomatic individuals, few of whom may be children.

We therefore used the second data set collected in this study to examine the secondary attack rate of children versus adults in household clusters where adults were identified as an index case. In 11 observational studies of household transmission clusters, the secondary attack rate in tested paediatric household contacts (<18 years old) was significantly lower than that in adult household contacts RR=0.62 (95% CI, 0.42-0.91) (Figure 3).

**Figure 3.**
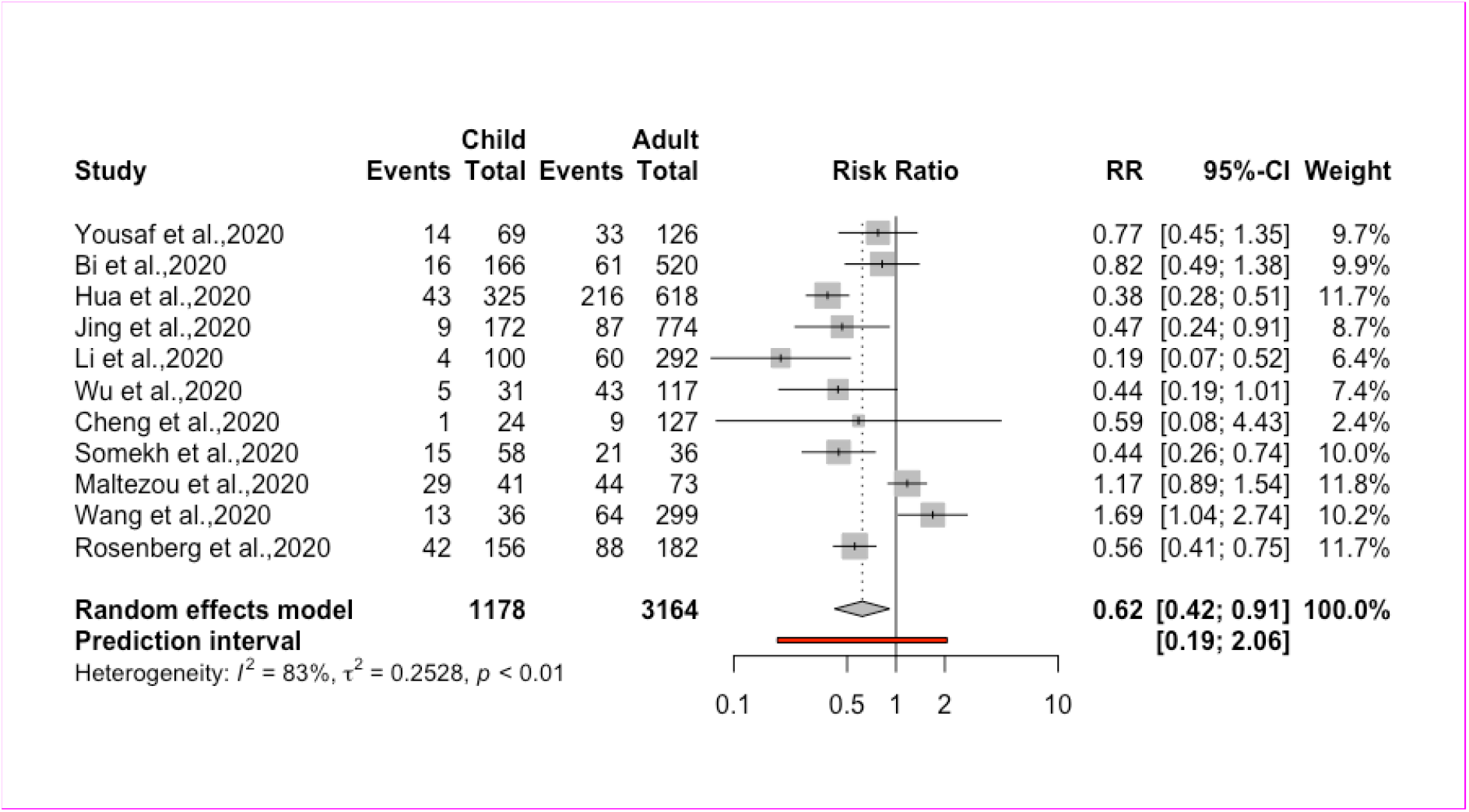
Relative risk (RR) for the secondary attack rate of children and adults in household SARS-CoV-2 transmission clusters. Events describe the number of SARS-CoV-2 positive individuals identified in the study.

In a subset analysis where additional information was provided on the age of the paediatric contact, younger children (<10 years) were no more or less susceptible to infection compared to older children (>10 years) RR = 0.69 (95% CI, 0.26-1.82) with no significant heterogeneity (I^2^=33%, P=0.17, Figure 4). Together, these data suggest that children (<18 years old) are less susceptible to SARS-CoV-2 infection in a household transmission cluster.

**Figure 4.**
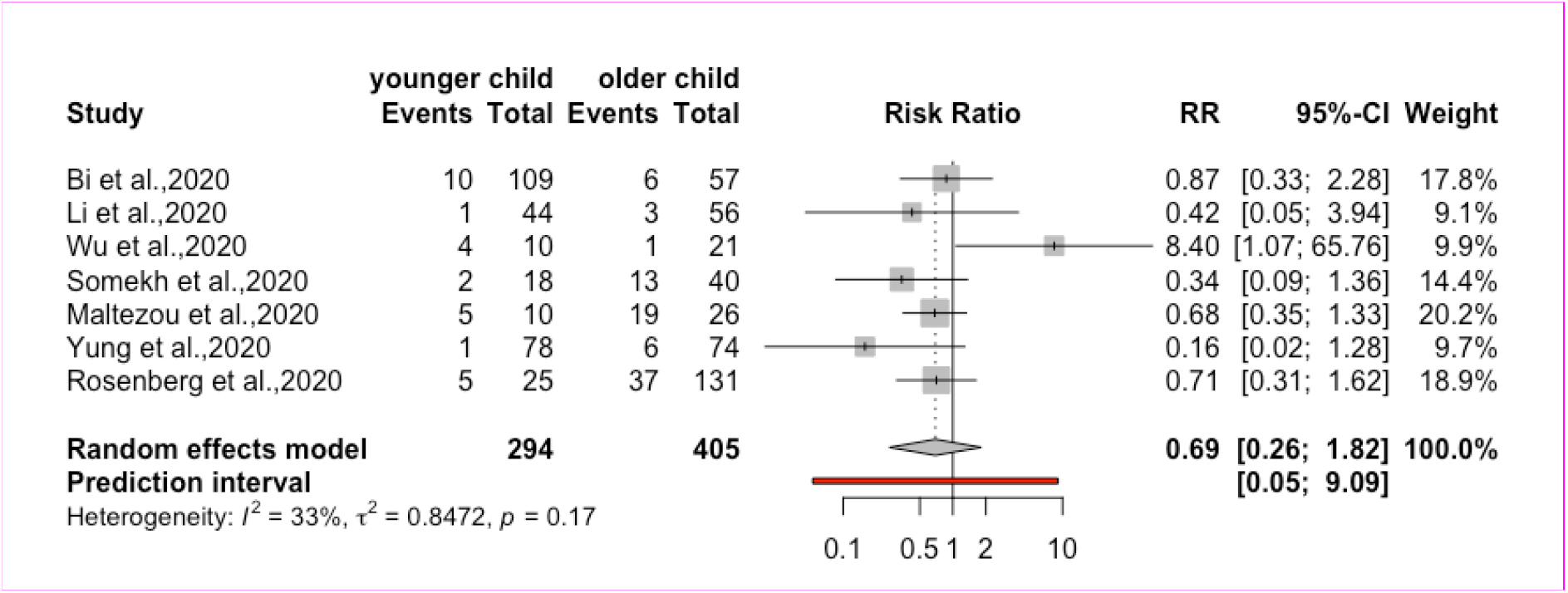
Relative risk (RR) for the secondary attack rate of younger and older children in household SARS-CoV-2 transmission clusters. Events describe the number of SARS-CoV-2 positive individuals identified in the study.

## DISCUSSION

The transmission of SARS-CoV-2 to and from children has remained controversial throughout the course of the COVID-19 pandemic. In the present study, we only recorded a limited number of household transmission clusters (3.8%) where children (<18 years) were identified as the index case. This observation is supported by previous study from China, where a study of 66 family transmission clusters showed that children were never the first in the family to be diagnosed with COVID-19 [83]. Other reviews have also found limited evidence of children as the index case in household transmission clusters [4, 20]. Indeed, in a family based study by the Dutch National Institute for Public Health and the Environment there were no indications in any of the 54 participating families that a child <12 years old was the source of COVID-19 within the family[84]. A more recent study of households in Tennessee and Wisconsin suggested that of 101 family SARS-CoV-2 clusters, only 14 had paediatric index cases [85].

There are multiple possible reasons as to why children may be infrequently identified as the index case in household transmission clusters. This may reflect limited interaction of children outside the home during the period in question or the higher probably of an adult traveling to a COVID-19 endemic area than a child (. An alternate hypothesis is that children are less susceptible to SARS-CoV-2 infection than adults. Indeed, this is consistent with our observation that in household transmission clusters, children were significantly less likelyto acquire SARS-CoV-2 than their adult family members. Interestingly, we found that older children were not significantly more likely than younger children to acquire the virus, in contrast to previous pre-print suggestions [4].

A reduced incidence of SARS-CoV-2 infection in children outside the home has previously been reported [2, 4, 8, 18]. [86]. [87]. Indeed, these data are congruent with survey data from Vo’, Italy [88]. Here, all age groups were homogenously sampled yet no children tested positive for SARS-CoV-2 infection. This was despite the fact that at least 13 of these children lived together with infected family members [88].

Once infected, it remains to be determined as to whether children are more or less likely to transmit the SARS-CoV-2 to a family member as an infected adult. Whilst the mean number of infected household members was lower when a child was identified as the index case of the cluster, the low number of clusters eligible for inclusion in this analysis precluded any definitive conclusions. However, these data are consistent with recent U.S. data suggesting that the SAR from paediatric index cases (18 years old) was less than that of adults (43% and 57% respectively) [86]. It has previously been suggested that children are less likely to transmit SARS-CoV-2 compared to adults [7, 8, 18-20]. Such suggestions have remained controversial amongst other findings that children have equivalent nasopharyngeal viral loads to adults [14, 16, 17]. However, reanalysis of the aforementioned studies have shown that i) young children (<10 years old) did indeed have a significantly lower viral load [19] ii) that the comparison was being performed between children in the first 2 days of symptoms and hospitalized adults with severe disease[17] and iii) datasets included few children younger than 16 years[16]. Similarly, pre-print suggestions that the risk of SARS-CoV-2 transmission to contacts is greatest from infected individuals <14 years old may be affected by limited case numbers in this age group (14/1489 total cases)[12].

Should children be less likely to transmit the virus, it is interesting to speculate the possible mechanism by which this occurs. There is an emerging body of evidence that mild or asymptomatic patients are less infectious than those with pronounced clinical symptoms [7, 13, 80, 81, 89]. Indeed, our meta-analysis showed that an asymptomatic index case was associated with a significantly lower secondary attack rate compared to a symptomatic index case. It is therefore tempting to hypothesise that children may be less infectious than adults infected with SARS-CoV-2 due to their more mild clinical manifestation of disease. However, such a hypothesis requires validation across a larger and more diverse dataset.

The present study was subject to several important limitations. Firstly, as we were conservative during the data collection only a limited number of studies were included, potentially contributing to the high I2 value observed. This study also assumes that SARS-CoV-2 infections in the household contacts of infected individual were the result of a direct transmission event. However, it is possible that the household contact acquired the virus from another source (e.g. from community exposure) and that the first in the family to develop symptoms was not necessarily the index case. We were also unable to control for the chance of a ‘common exposure’ where two individuals were infected by the same source at the same time, but one individual was incorrectly identified as the sole index case of the cluster as they were the first to develop symptoms. Indeed, this appears to have confounded the analysis of a series of family clusters identified in South Korea [5, 15]. It is also important to note that we were unable to differentiate between pre-symptomatic and asymptomatic infections and therefore the number of identified asymptomatically infected individuals may be overestimated. Finally, it is important to note that these data should not be extrapolated to SARS-CoV-2 transmission outside the home where children tend to make more social contacts than adults[18]. This could significantly influence transmission dynamics in the community setting, although our data are congruent with the low rate of SARS-CoV-2 transmission in Australian schools [90].

We are almost one year into the COVID-19 pandemic and many countries are still struggling to control outbreaks of SARS-CoV-2. At the time of writing, numerous countries in Europe have been forced into a second lockdown. However, unlike the first lockdown many countries (including the U.K. and Germany) have elected to keep schools open. The data presented in this manuscript suggest that should children become infected at school during this period, they are unlikely to spread SARS-CoV-2 to their co-habiting family members.

There is now a growing body of evidence that a safe and at least partially effective vaccine will be soon licenced for human use. However, due to the global demands, SARS-CoV-2 vaccines are likely to be first administered to pre-defined priority populations. Whilst prioritising the vaccination children against influenza has proved an effective tool in the reducing the spread of influenza virus in the community[10], our data suggest that a similar strategy would be unlikely to significantly decrease the household transmission of SARS-CoV-2.

## Data Availability

The authors have full access to all the data in the study.

## ACKNOWLEDGEMENTS

KRS was supported by the Australian Research Council [DE180100512]. ACB receives funding from the National Health and Medical Research Council with an Investigator Award (1175509).

